# Efficacy and Safety of Gene III L-ergothioneine Eye Care Solution in Dry Eye Syndrome: A Randomized, Self-Controlled Clinical Trial

**DOI:** 10.64898/2025.12.30.25343188

**Authors:** Rujun He, Jiang Jiang, Wei Ding, Feng Shao, Juan Cao, Xinyu Kuai, Hongying Ju, Guohua Xiao

## Abstract

**Purpose:** To evaluate the efficacy and safety of Gene III L-ergothioneine (EGT) Eye Care Solution for alleviating ocular discomfort in patients with dry eye syndrome (DES).

**Methods:** In this single-center, randomized, open-label, self-controlled trial, 40 DES patients were randomly assigned in a 1:1 ratio to one of two treatment groups: Group 1 received the Gene III EGT Eye Care Wash Solution via an eye wash cup (5 mL/vial), and Group 2 received the same formulation as Gene III EGT Eye Care Drops Solution (0.5 mL/vial). Outcomes were assessed at baseline and after 14 days using validated scales, including the Chinese Dry Eye Questionnaire, the Ocular Surface Disease Index (OSDI), and the Comprehensive Assessment of Visual Fatigue (CAVF), along with fluorescein tear film break-up time (TBUT).

**Results:** Combined analysis (N = 40) demonstrated significant improvements in dry eye symptoms with mean scores decreasing from 12.50 to 10.65 (P = 0.0353). Ocular Surface Disease Index (OSDI) scores improved from 12.25 to 9.2 (P = 0.0309), and visual fatigue, assessed by the CAVF scale, decreased from 11.18 to 6.60 (P = 0.0008). Significant increases in TBUT were observed in both groups, with left eye first TBUT rising in Group 1 (P = 0.0199) and Group 2 (P < 0.0001). No treatment-related adverse events were reported.

**Conclusion:** Gene III EGT Eye Care Solution effectively alleviated DES symptoms and demonstrated a favorable safety profile. Larger placebo-controlled trials are warranted to confirm these findings.

## Introduction

Dry eye syndrome (DES) is currently the most common ocular surface disorder in clinical ophthalmology and is characterized by a multifactorial etiology. Epidemiological data indicate that the global prevalence of DES ranges from approximately 5% to 50%. In China, epidemiological studies have reported a prevalence of about 21.0%–52.4% [1]. Accumulating evidence indicates that oxidative stress and inflammation are key drivers of DES pathogenesis [2-5].

To mitigate oxidative stress, various antioxidant-based interventions have been explored in clinical practice. Local therapeutic strategies commonly incorporate antioxidants such as coenzyme Q10 (CoQ10), vitamins A and E, and polyphenols (e.g., curcumin and epigallocatechin gallate) into artificial tear formulations to scavenge reactive oxygen species (ROS) and protect the corneal epithelium [6]. Systemic interventions primarily involve dietary supplementation with nutrients like omega-3 fatty acids and astaxanthin to attenuate ocular surface inflammation [7]. Despite providing symptomatic relief, these conventional therapies face several critical hurdles. Many natural antioxidants exhibit poor physicochemical stability and are highly susceptible to degradation by light, heat, and oxygen during storage and application [6]. In addition, therapeutic bioavailability is severely constrained by limited ocular bioavailability, owing to the corneal epithelial barrier and rapid nasolacrimal drainage, which hinder the maintenance of effective drug concentrations over time [8]. Crucially, most current antioxidants rely on passive diffusion for tissue entry and lack specific intracellular targeting mechanisms, preventing efficient delivery to mitochondria—the epicenter of ROS generation and oxidative damage [9]. Collectively, these limitations underscore the need for novel antioxidant agents with robust stability, enhanced bioavailability, and the capacity for targeted intracellular transport.

L-ergothioneine (EGT) is a natural amino acid first discovered by French pharmacist Charles Tanret in 1909 during his investigation of ergot fungi [6]. EGT is widely distributed across various human tissues and organs, with relatively high concentrations accumulating in those particularly susceptible to oxidative stress-induced damage, including the brain, eyes, liver, kidneys, bone marrow, and lymphoid tissues [7].

Studies have shown that EGT possesses potent antioxidant and anti-inflammatory effects [8, 9], and thus holds promise as a novel antioxidant for the treatment of AMD [10]. Increasing evidence also demonstrates a close and bidirectional relationship between oxidative stress and DES. Tear film instability triggers the production of reactive oxygen species (ROS) via the oxidative stress pathway; these ROS can damage the myelin sheath of ocular surface nerves and disrupt the homeostasis of the tear film lipid layer, thereby inducing or exacerbating ocular surface inflammation. Antioxidant therapy seeks to target the key pathogenic factors of DES, interrupt the vicious inflammatory cycle at various stages of the disease, and ultimately alleviate clinical symptoms [11]. Despite promising preclinical findings, clinical data on the efficacy of EGT in DES remain scarce. Therefore, this study aims to investigate the efficacy and safety of topical EGT administration in DES patients.

## Materials and Methods

### Materials

This prospective study was conducted as a single-center, randomized, open-label, parallel-group, and self-controlled clinical trial. The study enrolled patients with ocular discomfort who were treated at Hefei First People’s Hospital from November 4, 2024, to December 3, 2024. The study protocol comprised a screening period, a trial period, and a follow-up period. Efficacy evaluations were carried out at the end of the study, while safety evaluations were conducted throughout the study.

#### Inclusion Criteria

Participants were eligible for enrollment only if they met all the following criteria: (1) Provided written informed consent before screening; (2) Males aged 18–70 years or non-pregnant, non-lactating females; (3) Reported at least one of the following subjective symptoms: dry eyes, foreign body sensation, burning sensation, ocular fatigue, discomfort, redness, or fluctuating vision, with a score of ≥7 on the Chinese Dry Eye Questionnaire; (4) Had no plans for pregnancy, sperm donation, or egg donation within one month after the last use of the study product.

#### Exclusion Criteria

Participants were excluded if they met any of the following criteria: (1) Known contraindications or allergies to the study product; (2) Presence of active ocular infection, inflammation, or allergy in either eye during the screening period; (3) History of ocular herpes or current long-term oral antiviral therapy for herpes-related conditions; (4) Complete absence of congenital lacrimal or meibomian glands in the eyes; (5) Significant conjunctival scarring in the study eye(s); (6) Severe ocular disease, systemic disease, or uncontrolled medical conditions that may confound study evaluations or limit compliance; (7) History of incisional ocular surface surgery within one month before screening; (8) Use of other investigational drugs or devices within three months before screening; (9) Pregnant or lactating women, women with a positive pregnancy test, or those planning pregnancy; (10) Any condition deemed by the investigator to render the participant unsuitable for study participation.

A total of 40 participants were enrolled in this study, all of Han ethnicity, including 8 males and 32 females. The mean age was 27.1 ± 9.1 years, mean height 166.8 ± 8.0 cm, mean weight 61.9 ± 10.8 kg, and mean body mass index (BMI) 22.1 ± 2.7 kg/m^2^ (Table 1).

**Table 1.** Baseline Demographic and Clinical Characteristics.

| Parameter | Group 1 (N = 20 patients, 40 eyes) | Group 2 (N = 20 patients, 40 eyes) | Total (N = 40 patients, 80 eyes) |
| --- | --- | --- | --- |
| Sex (female:male), n | 17:3 | 15:5 | 32:8 |
| Age (years), mean ± standard deviation (SD) | $24.9 \pm 7.1$ | $24.7 \pm 8.0$ | $27.1 \pm 9.1$ |
| Height (cm), mean ± SD | $165.5 \pm 7.4$ | $165.6 \pm 8.7$ | $166.8 \pm 8.0$ |
| Weight (kg), mean ± SD | $61.0 \pm 9.8$ | $57.2 \pm 9.6$ | $61.9 \pm 10.8$ |
| BMI (kg/m<sup>2</sup>), mean ± SD | $22.2 \pm 3.0$ | $20.8 \pm 2.7$ | $22.1 \pm 2.7$ |
| Chinese Dry Eye Questionnaire Scale, mean | 12.40 | 12.60 | 12.50 |
| OSDI score, mean | 12.55 | 11.95 | 12.25 |
| CAVF score, mean | 11.50 | 10.85 | 11.18 |

Participants were randomly assigned in a 1:1 ratio to Experimental Group 1 or Experimental Group 2 using the block randomization method, with 20 subjects allocated to each group. Participants in Experimental Group 1 received L-ergothioneine Eye Care Solution administered via an eye wash cup (5 mL/vial). One tube was used per application, twice daily for 14 consecutive days. Participants in Experimental Group 2 received the same formulation administered as eye drops (0.5 mL/vial), with three drops per application (one tube), twice a day, for 14 consecutive days. Both formulations of Gene III L-ergothioneine Eye Care Solution were provided by Gene III Biotechnology Co., Ltd. The formulation contained purified water, 0.5% EGT, 0.05% sodium hyaluronate, taurine, and sodium chloride. This study was conducted in strict accordance with the ethical guidelines of the Declaration of Helsinki. The protocol was reviewed and approved by the Clinical Trial Ethics Committee of Hefei First People’s Hospital (Review No.: 2024-Lunshen-28). All procedures complied with Good Clinical Practice (GCP) guidelines, the Declaration of Helsinki, and applicable national laws and regulations. Written informed consent was obtained from all participants prior to enrollment, ensuring full protection of their rights and interests.

## Methods Study

### Design

This prospective, single-center, randomized, open-label, self-controlled trial (Ethics Approval No.: 2024-Lunshen-28) was conducted at Hefei First People’s Hospital between November and December 2024. A total of 40 DES patients were enrolled (mean age: 27.1 ± 9.1 years).

### Interventions

Participants were randomly assigned in a 1:1 ratio to one of two treatment groups:

Group 1 (N = 20): 5 mL/vial Gene III EGT Eye Care Solution (containing 0.5% EGT and sodium hyaluronate), administered via eye wash cups twice daily for 14 consecutive days.

Group 2 (N = 20): 0.5 mL/vial Gene III EGT Eye Care Solution of the same EGT formulation, administered as eye drops twice daily for 14 consecutive days.

### Outcome Measures

#### Primary Efficacy Endpoints

The primary efficacy endpoint was the change from baseline to Day 14 in the Ocular Surface Disease Index (OSDI) score. The OSDI is an internationally recognized, reliable, and validated tool for quantifying dry eye–related symptoms and their impact on vision-related quality of life [16].

#### Secondary Efficacy Endpoints

Secondary efficacy endpoints included: (1) Change from baseline to Day 14 in the Chinese Dry Eye Questionnaire (CDEQ) score, to assess symptom improvement specific to the Chinese population. (2) Change from baseline to Day 14 in the Comprehensive Assessment of Visual Fatigue (CAVF) scale score, to evaluate the impact of treatment on eye strain and visual fatigue. (3) Change from baseline to Day 14 in fluorescein tear break-up time (TBUT), an objective indicator of tear film stability.

In addition, the incidence of adverse events (AEs) was recorded throughout the treatment period to support the overall evaluation of treatment safety.

### Safety Evaluation Endpoints

The safety profile of the investigational Gene III EGT Eye Care Solution was comprehensively assessed in accordance with the Common Terminology Criteria for Adverse Events (CTCAE), v5.0. Slit-lamp examinations were performed at enrollment, during treatment (Day 8), and at study completion (Day 14). The cornea was systematically evaluated for corneal edema, epithelial microcysts, neovascularization, infiltrates, and fluorescein staining. The anterior segment, including the anterior chamber, iris, pupillary morphology, and lens clarity, was also examined to detect any abnormal changes. Adverse events occurring during the trial were collected using standardized questionnaires.

### Sample Size Estimation

This study was designed as a preliminary, exploratory trial to assess the safety and explore efficacy trends of the investigational Gene III EGT Eye Care Solution in patients with DES. At the study design stage, the sample size was determined pragmatically based on feasibility considerations and sample sizes commonly adopted in prior similar interventional studies for dry eye disease. Accordingly, a total of 40 subjects was pre-determined for enrollment. According to relevant expert consensus guidelines [17], this sample size is deemed adequate for detecting clinically meaningful changes in the primary efficacy endpoint in exploratory settings. Formal sample size calculations will be performed in future confirmatory trials using effect estimates derived from the findings of this exploratory study.

### Statistical Methods

#### Analysis of Efficacy Evaluation Indicators

All statistical analyses were performed using SAS software (version 9.4). Within-group comparisons between post-treatment visits and baseline were performed using paired t-tests when the data met assumptions of normality and homogeneity of variance. When these assumptions were not satisfied, the rank-sum test was applied. Between-group comparisons at the same post-treatment visit were analyzed using independent t-tests for normally distributed data with equal variances; otherwise, the rank-sum test was used. A significance level of P < 0.05 was considered statistically significant.

#### Analysis of Safety Evaluation Indicators

Safety analyses were performed using SAS software (version 9.4). All adverse events were described according to treatment group. The incidence, severity, and correlations between adverse events and the study product were recorded and analyzed. Safety was monitored continuously throughout the study period.

Primary endpoints included changes in symptom scores (CDEQ, OSDI, CAVF, and TBUT).

## Results

### Results of Efficacy Evaluation Indicators

#### Chinese Dry Eye Questionnaire Scale (CDEQ) Scores

Both treatment groups demonstrated a reduction in CDEQ scores at Day 14 compared with baseline. In Group 1 (5 mL Gene III EGT Eye Care Wash Solution), the mean score decreased from 12.40 at baseline to 10.65 at the end of the trial. In Group 2 (0.5 mL Gene III EGT Eye Care Drops Solution), the mean score decreased from 12.60 to 10.65. After combining the data from two groups, the mean score decreased from 12.50 at baseline to 10.65 at Day 14. This reduction was statistically significant (P = 0.0353), indicating that the study products effectively alleviated subjective dry eye symptoms (Figure 1).

**Figure 1.**
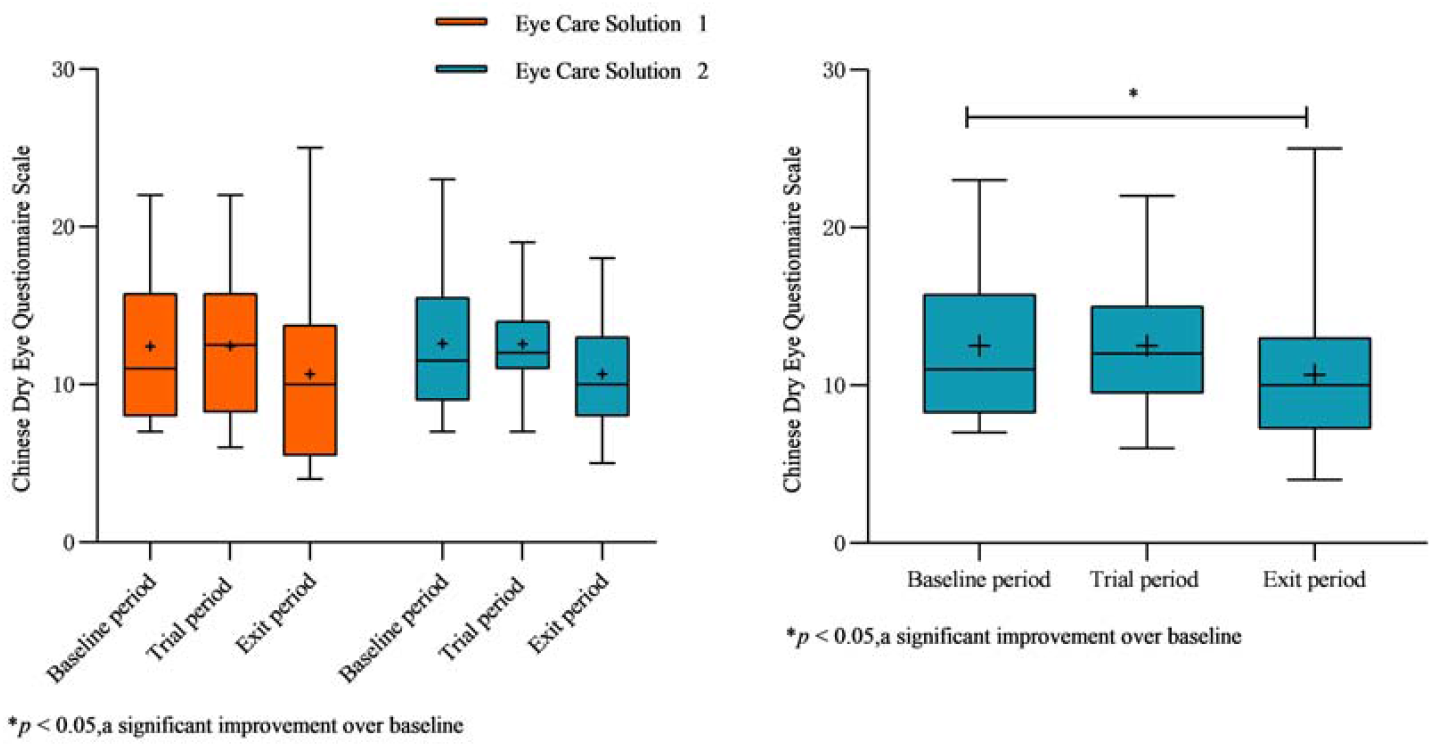
Box plots of CDEQ scores before (baseline, left) and after treatment (Day 14, right) in both groups. A significant decrease was observed after using the study products (P = 0.0353).

### Ocular Surface Disease Index (OSDI) Scores

Both groups showed reductions in OSDI scores at Day 14 compared with baseline. In Group 1, the mean OSDI score decreased from 12.55 at baseline to 10.00 at Day 14. In Group 2, the mean OSDI score decreased from 11.95 to 8.45. After combining the data from two groups, the reduction in OSDI scores was statistically significant (P = 0.0309) (Figure 2).

**Figure 2.**
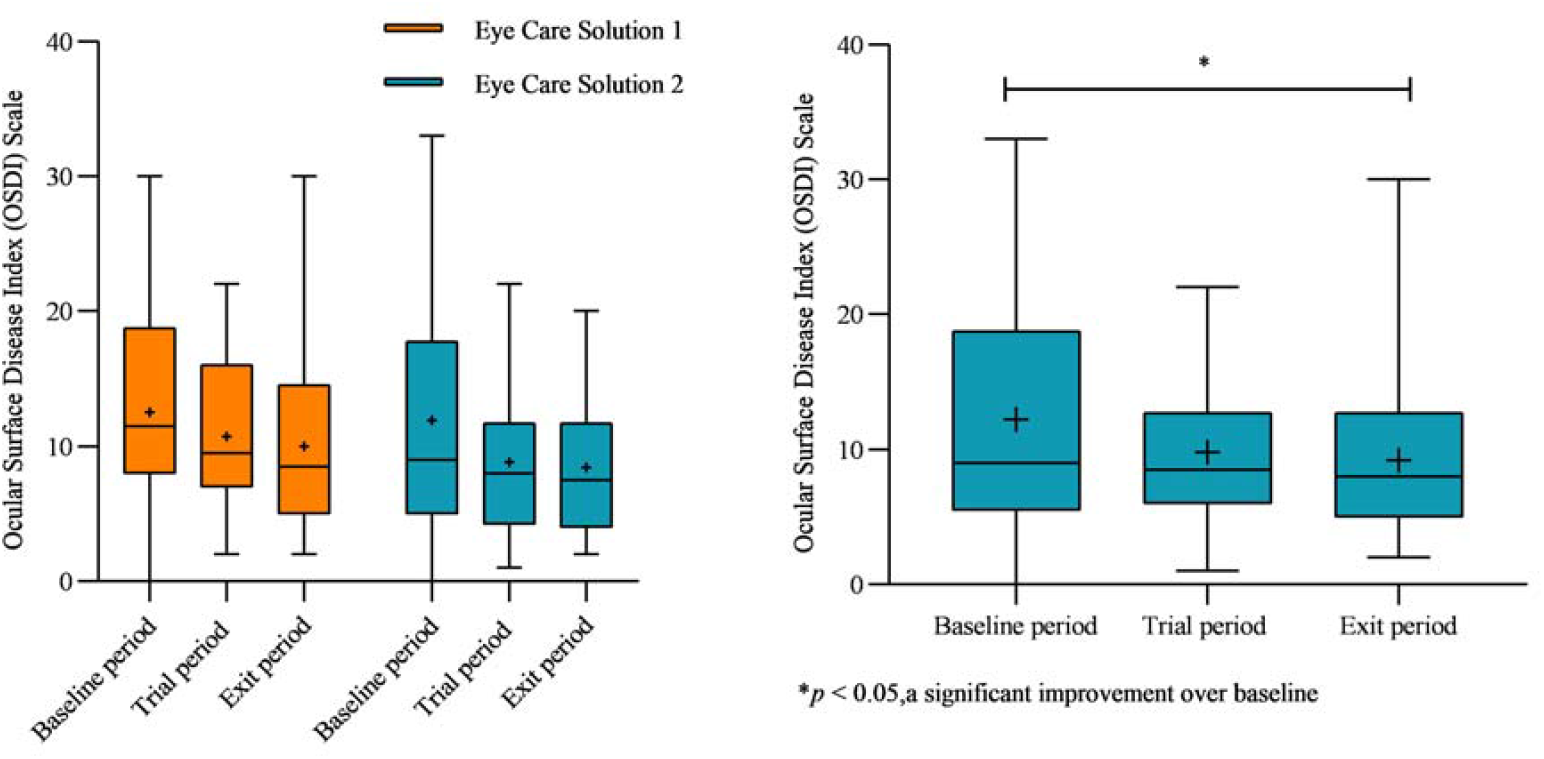
Box plots of OSDI scores before (baseline, left) and after treatment (Day 14, right) in both groups. A significant decrease was observed after using the study products (P = 0.0309).

### Visual Fatigue (CAVF) Scores

Both groups showed reductions in CAVF scores at Day 14 compared with baseline. In Group 1, the mean CAVF scale score decreased from 11.50 at baseline to 7.00 at Day 14. In Group 2, the mean CAVF scale score decreased from 10.85 to 6.20. After combining the data from two groups, the reduction in CAVF scores was statistically significant (P = 0.0008) (Figure 3).

**Figure 3.**
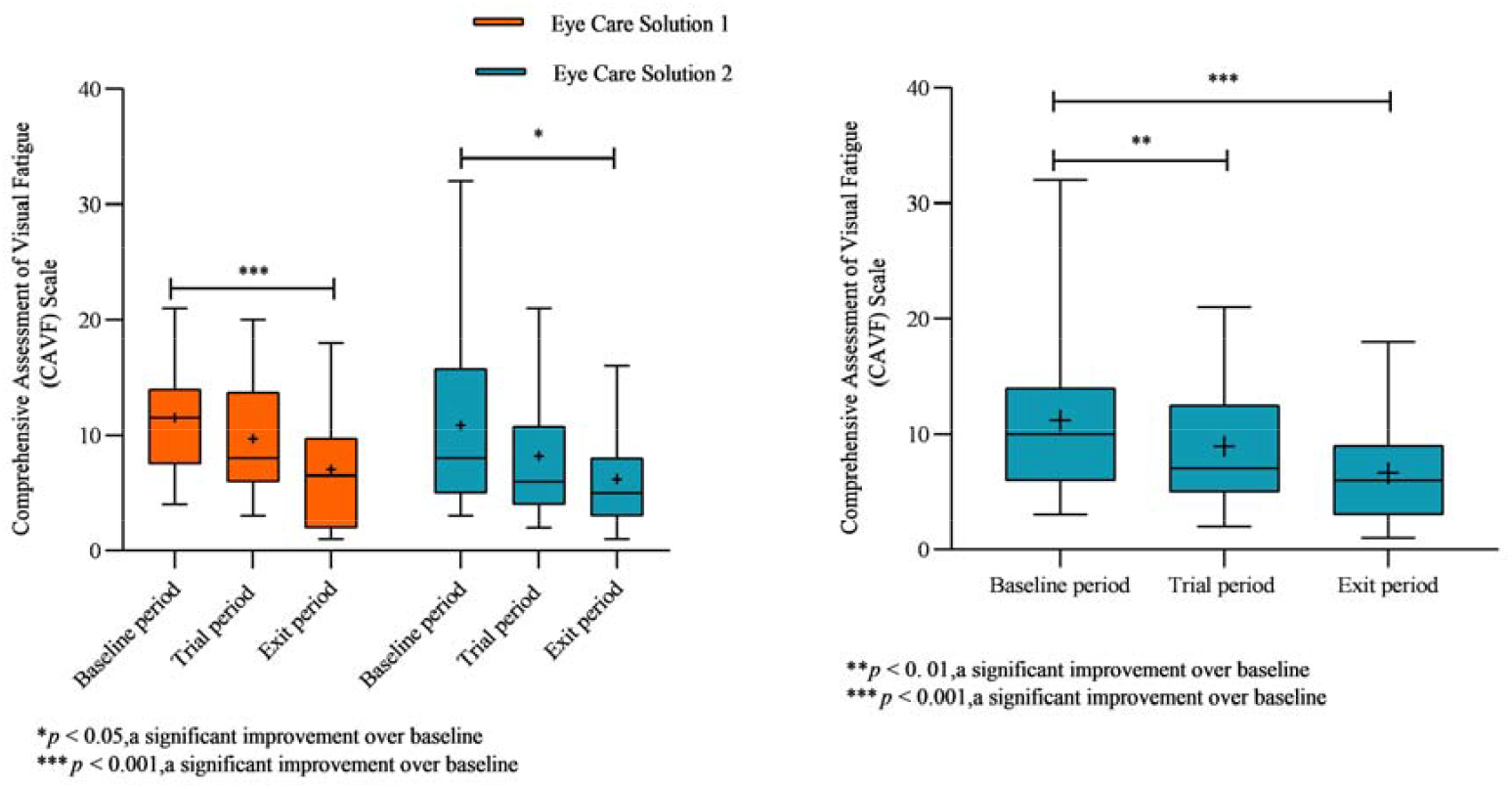
Box plots of CAVF scores before (baseline, left) and after treatment (Day 14, right) in both groups. A significant decrease was observed after using the study products (P = 0.0008).

### Tear Break-Up Time (TBUT) Measured by Fluorescein Staining

In Group 1, both the first and the average TBUT of the left eye and right eyes were significantly prolonged at Day 14 compared with baseline (Left eye: first TBUT P = 0.0199, average TBUT P = 0.0135; Right eye: first TBUT P = 0.0471, average TBUT P = 0.0222). In Group 2, similar significant prolongation was observed for both eyes (Left eye: first TBUT P = 0.0006, average TBUT P = 0.0003; Right eye: first TBUT P < 0.0001, average TBUT P < 0.0001) (Figures 4 and 5).

**Figure 4.**
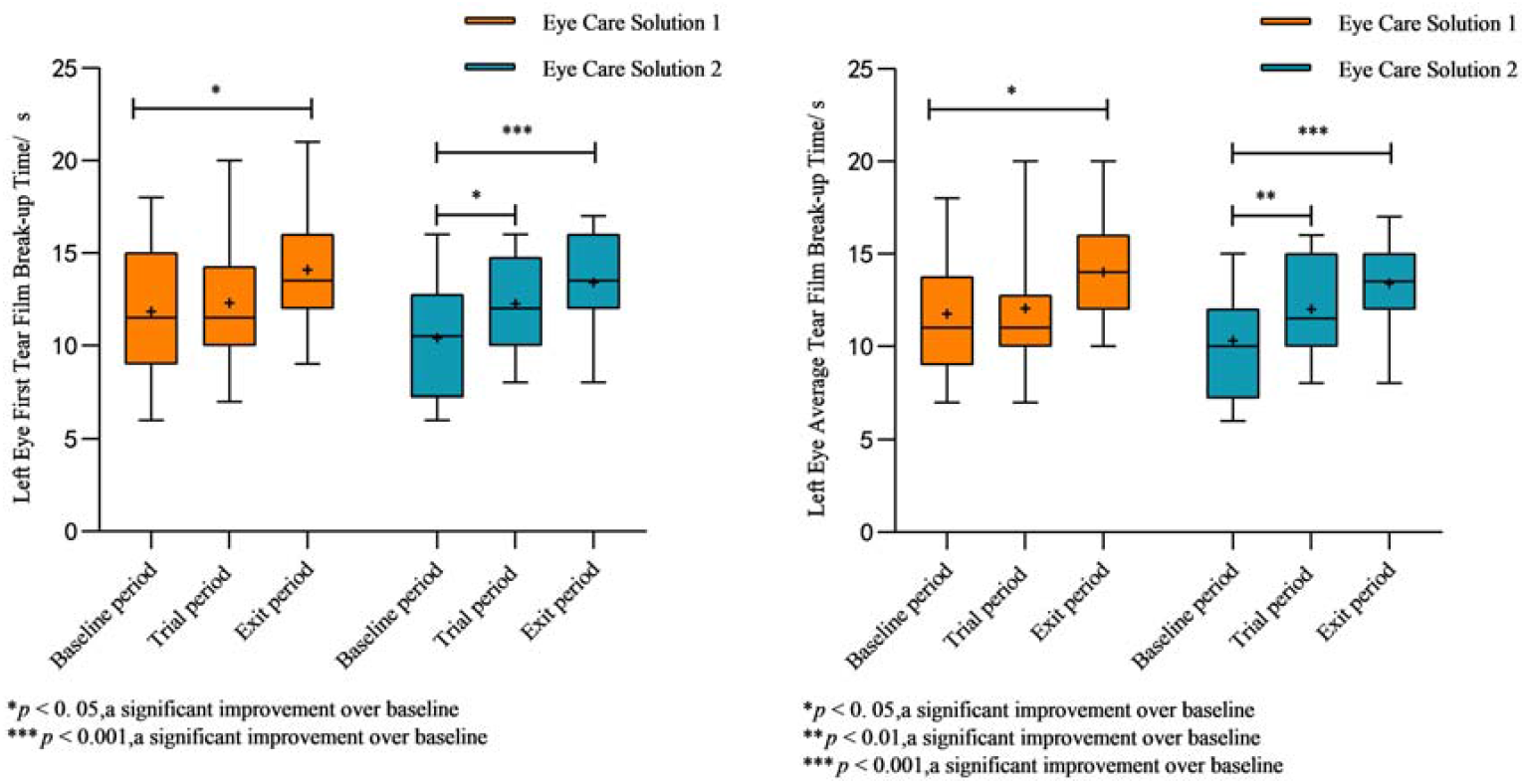
Box plots of first (left) and average (right) fluorescence-stained TBUT for the left eye in both groups. The average TBUT increased significantly after using the study products (P = 0.0199, 0.0135, 0.0471, 0.0222, respectively).

**Figure 5.**
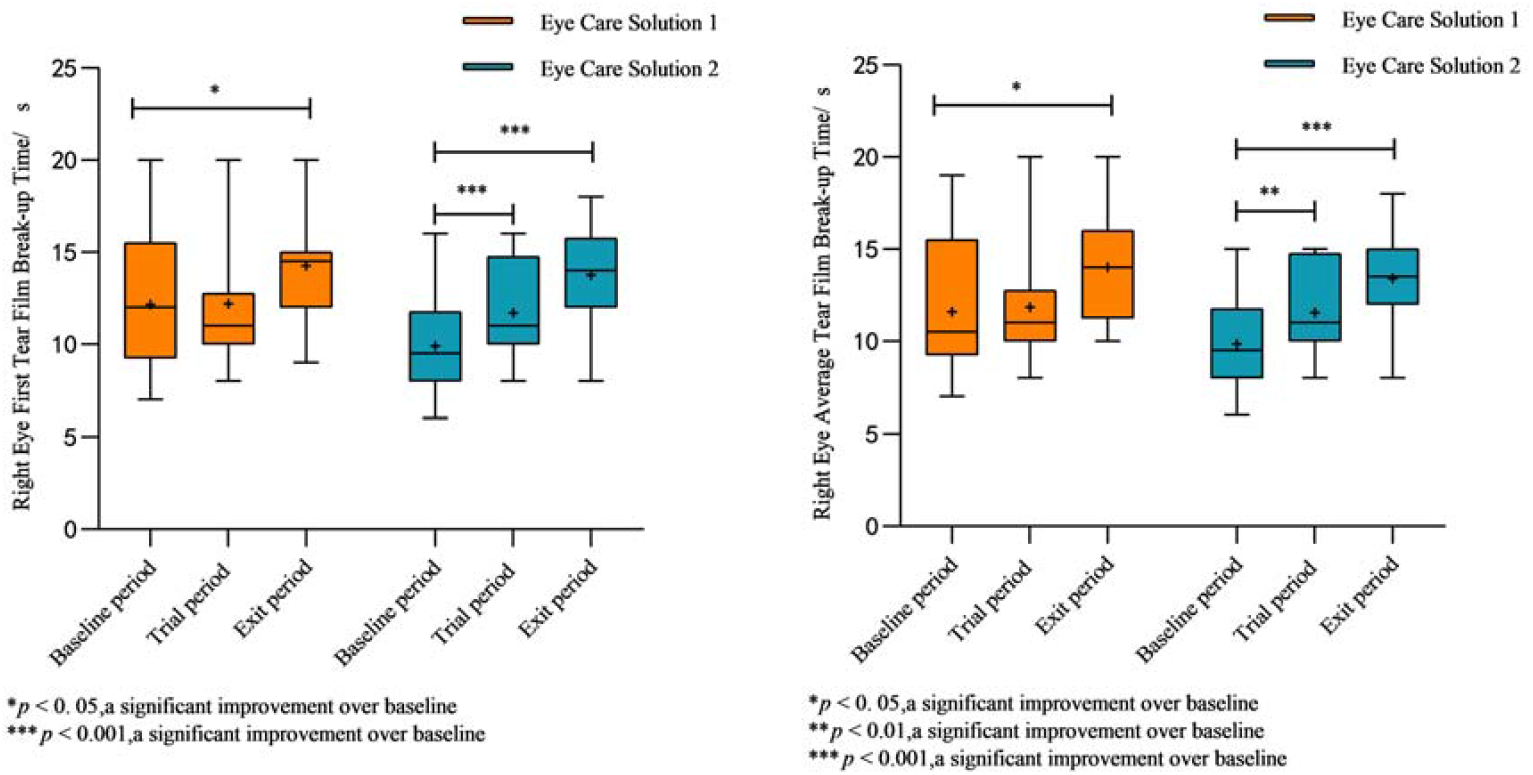
Box plots of first (left) and average (right) fluorescence-stained TBUT for the right eye in both groups. The average TBUT increased significantly after using the study products (P = 0.0006, 0.0003, < 0.0001, < 0.0001, respectively).

### Safety Assessment Results

A total of 40 subjects were recruited for this study. 20 subjects received 5 mL/vial of the Gene III EGT Eye Care Solution and 20 subjects received 0.5 mL/vial of the Gene III EGT Eye Care Solution. All subjects were included in the safety analysis group. No adverse events related to the study product were reported during the trial.

1. Corneal Safety: Assessment of corneal edema, epithelial microcysts, neovascularization, infiltration, and fluorescein staining at enrollment, Day□8, and Day□14 showed grade 0 in all subjects. No pathological changes such as corneal erosions, peripheral ulcers, or infected ulcers were observed, indicating that corneal structure and epithelial integrity were unaffected by the intervention.
2. Intraocular Structures: Slit-lamp examinations showed normal intraocular structures in all subjects. The anterior chamber depth and aqueous humour were normal and clear; iris morphology was intact; pupils were normal in size, shape, and light reflex; and the lens was clear with no opacities or morphological abnormalities. These findings confirm that intraocular structures were unaffected by the intervention.
3. Incidence of Adverse Events: Data collected through standardized questionnaires indicated that no adverse events (including eye irritation, pain, itching, burning, and other related symptoms) occurred.

## Discussion

Numerous studies have demonstrated that the absorption and transport of EGT are mediated by the specific organic cation transporters novel-1 (OCTN-1), encoded by the SLC22A4 gene [12-14]. OCTN-1 is closely related to inflammation regulation [15], and its widespread expression allows EGT to distribute across various organs, including the ocular surface, which is particularly susceptible to oxidative stress and inflammatory damage. Mitochondria are the primary site of intracellular free radical generation. EGT is transported into mitochondria through OCTN-1, where it effectively scavenges free radicals and mitigates mitochondrial oxidative damage [16]. According to the Consensus of Clinical Diagnosis and Treatment Experts for Dry Eye in China (2024), antioxidant eye drops such as Visomitin (SkQ1) have been employed in European and American countries to treat DES, providing a novel therapeutic option. SkQ1 targets mitochondria to neutralize oxidative free radicals, suppress ocular oxidative stress, and alleviate dry eye symptoms. In addition to reducing inflammation, it addresses tissue degeneration and tear film deficiencies. Phase II clinical trials in the United States have shown that SkQ1 effectively improves both symptoms and signs of dry eye. The mechanism of action of EGT closely parallels that of SkQ1, highlighting its great potential as a therapeutic agent for DES.

The ophthalmic solution used in this study contained 0.5% EGT and 0.05% sodium hyaluronate. Results demonstrated that the solution effectively alleviated dry eye symptoms, reduced signs of ocular surface disease, and relieved visual fatigue, indicating notable antioxidant and anti-inflammatory effects. In this study, 40 subjects were randomly enrolled: 20 subjects in Experimental Group 1 received 5 mL/vial of the solution administered via an eye wash cup, while 20 subjects in Experimental Group 2 received 0.5 mL/vial administered as eye drops. All subjects completed the study without adverse events and were included in the analysis.

Both groups showed a decreasing trend in primary symptom scales (CDEQ and OSDI), and the combined analysis reached statistical significance (P = 0.0353 and P = 0.0309, respectively). However, no significant difference was observed between the two groups. This suggests similarities in local bioavailability or the mechanisms of action between the two administration methods. While eye wash cup delivery may cover a larger ocular surface area, the drug residence time might be relatively short. In contrast, repeated eye drop instillations may maintain a more consistent drug concentration on the ocular surface. Both formulations used contained 0.05% sodium hyaluronate, whose moisturizing and retention properties likely mitigated differences between administration routes, enabling effective delivery of EGT to ocular surface tissues and exerting antioxidant and anti-inflammatory effects. Future studies could further evaluate the pharmacokinetic profiles of EGT on the ocular surface for these two administration methods, for example, by measuring drug concentrations in tear fluid or using fluorescence imaging techniques.

Regarding visual fatigue (CAVF) and tear film break-up time (TBUT), both within-group comparisons and combined analysis showed significant improvements (all P < 0.05), indicating that L-ergothioneine Eye Care Solution effectively alleviated dry eye-related visual fatigue and enhanced tear film stability, regardless of whether administered via eye wash cup or eye drops. The absence of adverse events in all subjects suggests a favorable safety profile for both administration methods.

Hiroyuki et al. administered eye drops containing anti-inflammatory ingredients, such as dipotassium glycyrrhizinate, to healthy subjects [24]. They observed a statistically significant improvement in DEQS scores after use (P < 0.001), but no significant changes were found in TBUT or other scores (P > 0.05). These findings suggest that the Gene III EGT Eye Care Solution holds certain advantages over similar products.

In conclusion, L-ergothioneine Eye Care Solution effectively alleviates dry eye symptoms, reduces ocular surface diseases, and relieves visual fatigue, while demonstrating a favorable safety profile.

The lack of significant difference in efficacy between the two administration methods may be attributed to the inclusion of sodium hyaluronate in the formulation, which prolongs ocular surface retention, along with the inherent good tissue permeability of EGT. These findings provide preliminary support for flexible clinical selection of administration routes, and further pharmacokinetic studies could offer more detailed mechanistic insights.

## Limitations

(1) This study adopted a self-controlled design. Although this approach reduces the influence of individual differences, it cannot fully eliminate time-related or regression effects. Future studies should consider placebo-controlled or active-controlled designs to more accurately evaluate the treatment effect. (2) The sample size was relatively small, which limits the statistical power and generalizability of the results. Subsequent studies should include larger cohorts to improve the external validity and reliability of the results. (3) The evaluation primarily relied on subjective scales, with limited objective indicators. Although these assessments reflect participants’ experiences and symptom changes, they may not fully capture all potential effects of the treatment. Future research should incorporate additional objective measurement tools and biomarker analyses to enhance the objectivity and comprehensiveness of the results.

## Data Availability

All data produced in the present work are contained in the manuscript

## Availability of Data

The datasets generated and analyzed during the current study are available from the corresponding author, [Author Jiang], upon reasonable request.

## Ethical Statement

The study protocol was approved by the Ethics Committee of Hefei First People’s Hospital (2024-Lunshen-28). Written informed consent was obtained from all participants prior to enrollment.

## Conflict of Interest

The authors declare no competing financial or non-financial interests.

## Acknowledgements

We thank Gene III Biotechnology Co., Ltd. for funding this work. We also appreciate the assistance of Anhui Wanbang Pharmaceutical Technology Co., Ltd. with resource coordination and Hefei First People’s Hospital for facilitating the clinical trials.

## Notes

### Competing Interest Statement

The authors have declared no competing interest.

### Clinical Trial

ChiCTR2400090987

### Funding Statement

This study did not receive any funding

### Author Declarations

The protocol was reviewed and approved by the Clinical Trial Ethics Committee of Hefei First People's Hospital (Review No.: 2024-Lunshen-28)

